# Economic Burden of Viral Acute Respiratory Infections in Upper-Middle-Income Countries: Protocol For A Systematic Review

**DOI:** 10.1101/2020.12.14.20248198

**Authors:** César Ramos Rocha-Filho, Aline Pereira da Rocha, Felipe Sebastião de Assis Reis, Ana Carolina Pereira Nunes Pinto, Gabriel Sodré Ramalho, Giulia Fernandes Moça Trevisani, Laura Jantsch Ferla, Maria Eduarda Santos Puga, Luci Correa, Virgínia Fernandes Moça Trevisani, Márcia Mello Costa De Liberal, Álvaro Nagib Atallah

## Abstract

**Objective:** To synthesize the available data on the economic burden of Coronavirus Diseases 2019 (COVID-19), Severe Acute Respiratory Syndrome (SARS), Middle East Respiratory Syndrome (MERS), Influenza-Like Illness (ILI), Respiratory Syncytial Virus (RSV)-related Acute Respiratory Infection (ARI), and Parainfluenza Virus type III (PIV3)-related ARI in Upper-Middle-Income Countries (UMIC), highlighting its major causes and comparing direct costs among nations.

**Study design:** Systematic review, following the recommendations proposed in the Cochrane Handbook, but with some adaptations from previous economic studies.

**Review question:** Is there any economic cost of viral ARI in UMIC?

**Types of studies to be included:** Partial economic evaluation, such as Cost-of-Illness (COI) studies and burden of illness/diseases, database analysis, observational reports (cross-sectional studies, and prospective and retrospective cohort), and economic modelling studies that discuss one of the viral ARI in UMIC. No year of publication filter or language limit will be applied.

**Search databases:** MEDLINE, EMBASE, LILACS, CINAHL, EconLit, CRD Library, MedRxiv, and Research Square. Moreover, hand searches of the bibliographies of included studies and relevant reviews identified during the screening process will be undertaken to identify any additional relevant study for inclusion in our review.

**Synthesis of results:** Qualitative analysis. We will focus on the overall economic burden of the diseases on health systems and population; total direct cost; the contribution of different cost components to the economic burden (e.g. pharmacological therapy, hospitalization); comparative assessments of costs analysis across geographical location and time horizon; and current research gaps. Moreover, we intend to identify, when presented, prevalence and incidence rates of each disease.

**PROSPERO registration number:** CRD42020225757.

## Background

Acute Respiratory Infections (ARI) are a worldwide threat responsible for more than four million deaths annually [1]. The pathogenicity is typically caused by a virus, bacteria, fungi, or a mix of these microorganisms [2]. Although knowledge of modes of transmission is ever-evolving, scientific consensus reports airborne transmission by droplets and contact as the main acquisition routes of most respiratory infection diseases, especially viral ARI [1,2].

According to the World Lung Foundation, mortality rates of viral ARI are particularly high in the youngest and oldest people in low- and middle-income nations [1]. In this context, the geographical setting presents itself as a host factor due to the inadequate availability and effectiveness of medical care and infection prevention, and control measures of these countries to contain spread, such as vaccines, access to health-care facilities, and capacity of isolation at the hospitals and outpatient settings (or home settings) [2].

Due to the high and increasing incidence and prevalence rates, viral ARI represents an enormous economic burden in healthcare systems, independently of income classification [1–3]. To contextualize, an economic burden of diseases is related to the financial loss of individuals, families, societies, and governments due to economic resources spent related to respond to the diseases [3,4].

Outbreaks such as Severe Acute Respiratory Syndrome (SARS) in 2003 [5], and more recently the Coronavirus Disease 2019 (COVID-19) pandemic in December 2019 [6], have shown the burden that viral ARI can inflict in healthcare systems and national economies. In this scenario, valid estimates of the Cost-of-Illness (COI) in different perspectives are of particular importance and necessity to justify preventive and intervention-based programs. In the same way, estimates of the overall health system economic impact of viral ARI help to understand what specific factors are driving these costs, and may inform decision-makers to support formulation and refinement of policies [3,4,7,8].

### Objectives

Our aim is to synthesize the available data on the economic burden of six viral ARI in Upper-Middle-Income Countries (UMIC), highlighting its major causes and comparing direct costs among nations.

## Methods

### Study design and registration

The synthesize process will be based on the methodological process of a systematic review. To the best of our knowledge, there is no formal guideline to perform a systematic review of economic evaluations [4,7,9]. Thus, we will follow the well-established recommendations proposed in the Cochrane Handbook [10], but with some adaptations from studies performed by previously authors [4,7,9,11,12]. The main modifications included quality assessment and data analysis process.

This protocol is registered in the PROSPERO ‘‘International Prospective Register of Systematic Reviews’’ (CRD42020225757) and structured following the Preferred Reporting Items for Systematic Review and Meta-Analysis (PRISMA) Protocols guidance [13].

### Definition of the research question

The research question was structured in an iterative way to reflect the critical points related to the economic burden of six viral ARI in UMIC. We accepted a decision-maker perspective and pursued to identify comparative indicators.

Thus, our systematic review will aim to answer the following research question: Is there any economic cost of viral ARI in UMIC? Moreover, we intend to identify the major costs drivers related to viral ARI in different countries with sociodemographic similarities.

### Criteria for considering studies for this review

A study will be eligible for inclusion if it presents a burden estimation, in monetary units, of one of the choose viral ARI in an UMIC. Thus, our systematic review will present three main “sets” of interest: economic evidence, viral ARI and UMIC. Table 1 details the definitions that will be adopted.

**Table 1.**
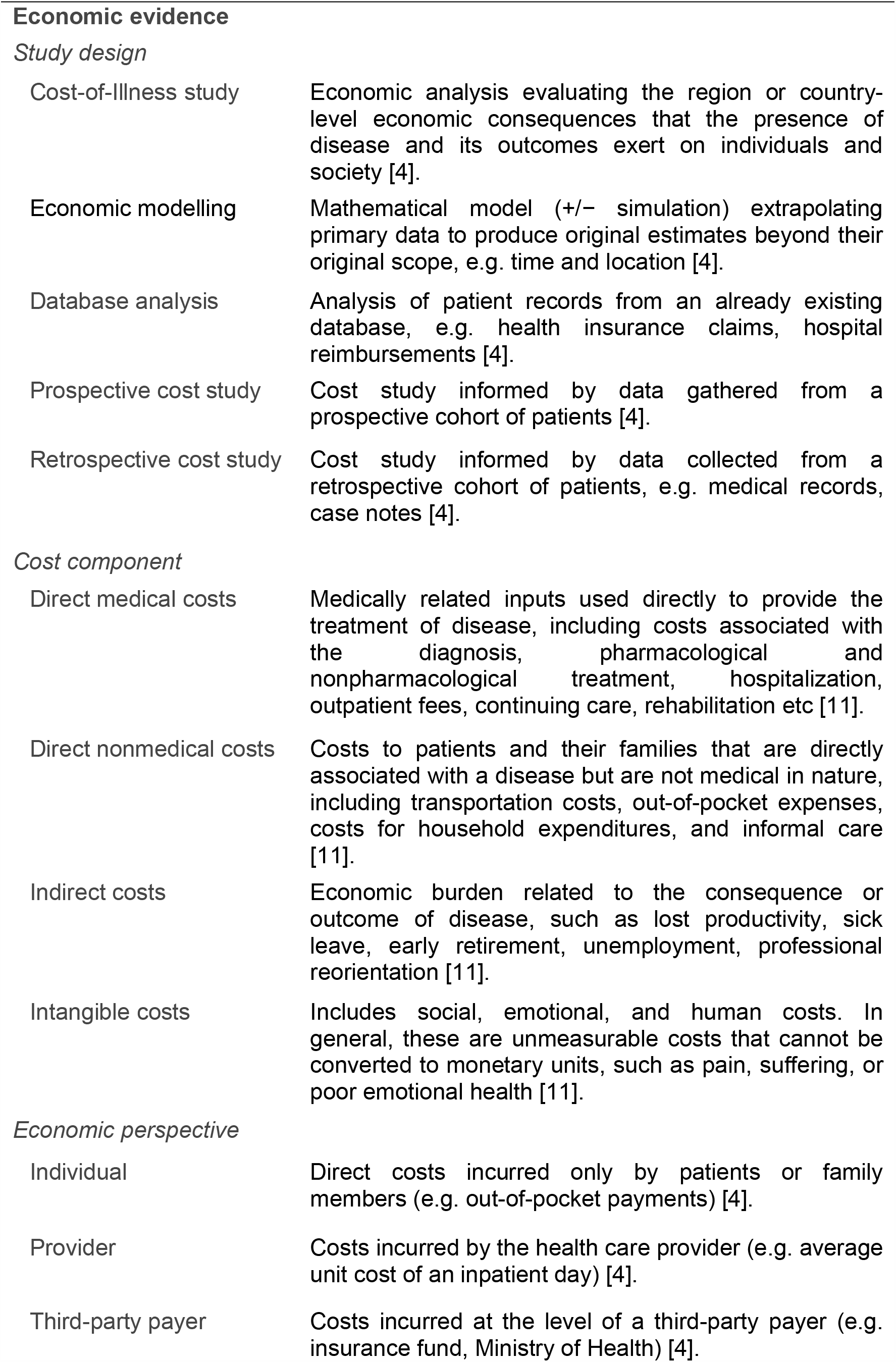
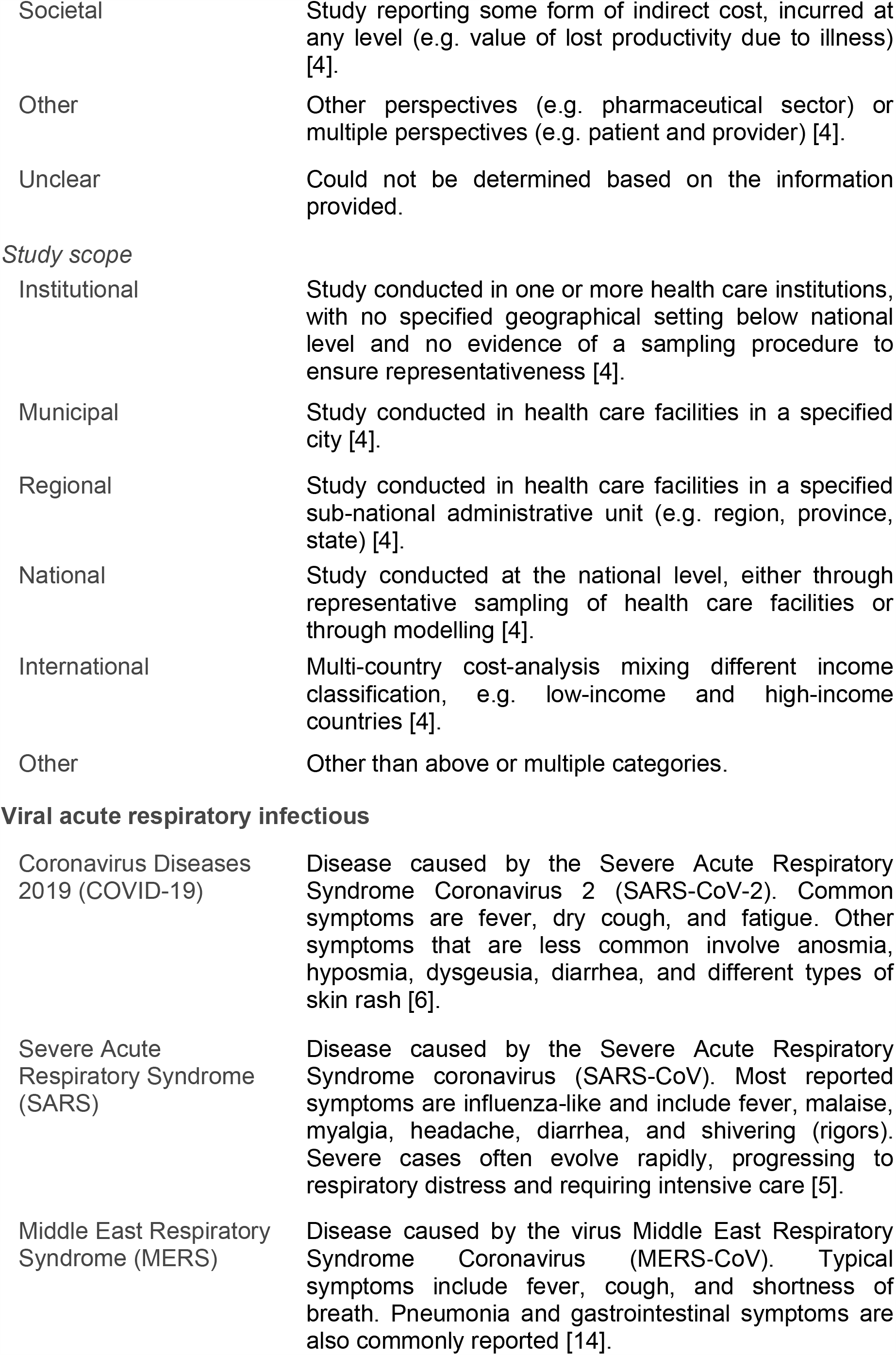
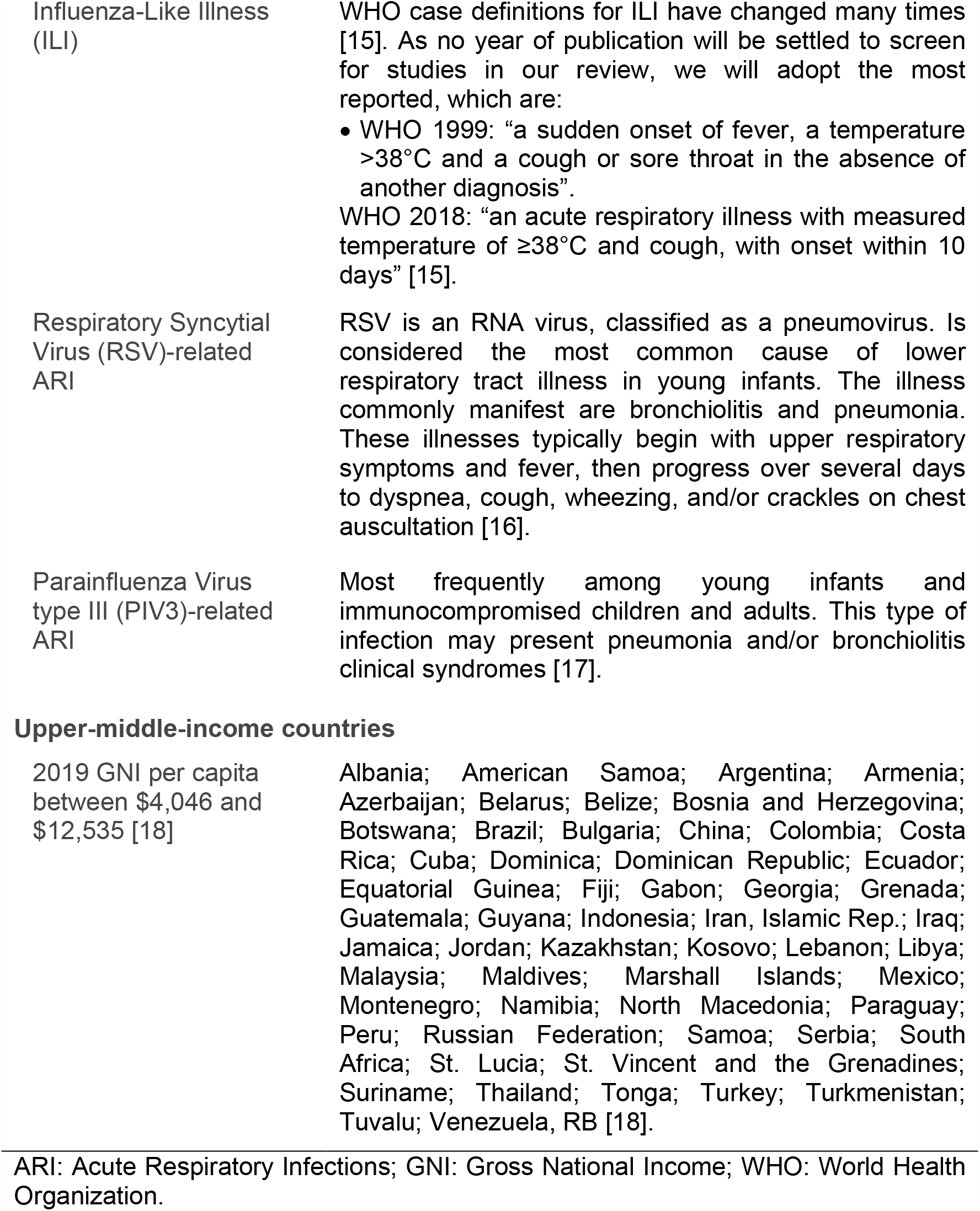
Definitions adopted in this systematic review to select and categorize studies.

| **Economic evidence** |  |
| --- | --- |
| *Study design* |  |
| Cost-of-Illness study | Economic analysis evaluating the region or country-level economic consequences that the presence of disease and its outcomes exert on individuals and society [4]. |
| Economic modelling | Mathematical model (+/- simulation) extrapolating primary data to produce original estimates beyond their original scope, e.g. time and location [4]. |
| Database analysis | Analysis of patient records from an already existing database, e.g. health insurance claims, hospital reimbursements [4]. |
| Prospective cost study | Cost study informed by data gathered from a prospective cohort of patients [4]. |
| Retrospective cost study | Cost study informed by data collected from a retrospective cohort of patients, e.g. medical records, case notes [4]. |
| *Cost component* |  |
| Direct medical costs | Medically related inputs used directly to provide the treatment of disease, including costs associated with the diagnosis, pharmacological and nonpharmacological treatment, hospitalization, outpatient fees, continuing care, rehabilitation etc [11]. |
| Direct nonmedical costs | Costs to patients and their families that are directly associated with a disease but are not medical in nature, including transportation costs, out-of-pocket expenses, costs for household expenditures, and informal care [11]. |
| Indirect costs | Economic burden related to the consequence or outcome of disease, such as lost productivity, sick leave, early retirement, unemployment, professional reorientation [11]. |
| Intangible costs | Includes social, emotional, and human costs. In general, these are unmeasurable costs that cannot be converted to monetary units, such as pain, suffering, or poor emotional health [11]. |
| *Economic perspective* |  |
| Individual | Direct costs incurred only by patients or family members (e.g. out-of-pocket payments) [4]. |
| Provider | Costs incurred by the health care provider (e.g. average unit cost of an inpatient day) [4]. |
| Third-party payer | Costs incurred at the level of a third-party payer (e.g. insurance fund, Ministry of Health) [4]. |
| Societal | Study reporting some form of indirect cost, incurred at any level (e.g. value of lost productivity due to illness) [4]. |
| Other | Other perspectives (e.g. pharmaceutical sector) or multiple perspectives (e.g. patient and provider) [4]. |
| Unclear | Could not be determined based on the information provided. |
| *Study scope* |  |
| Institutional | Study conducted in one or more health care institutions, with no specified geographical setting below national level and no evidence of a sampling procedure to ensure representativeness [4]. |
| Municipal | Study conducted in health care facilities in a specified city [4]. |
| Regional | Study conducted in health care facilities in a specified sub-national administrative unit (e.g. region, province, state) [4]. |
| National | Study conducted at the national level, either through representative sampling of health care facilities or through modelling [4]. |
| International | Multi-country cost-analysis mixing different income classification, e.g. low-income and high-income countries [4]. |
| Other | Other than above or multiple categories. |
| **Viral acute respiratory infectious** |  |
| Coronavirus Diseases 2019 (COVID-19) | Disease caused by the Severe Acute Respiratory Syndrome Coronavirus 2 (SARS-CoV-2). Common symptoms are fever, dry cough, and fatigue. Other symptoms that are less common involve anosmia, hyposmia, dysgeusia, diarrhea, and different types of skin rash [6]. |
| Severe Acute Respiratory Syndrome (SARS) | Disease caused by the Severe Acute Respiratory Syndrome coronavirus (SARS-CoV). Most reported symptoms are influenza-like and include fever, malaise, myalgia, headache, diarrhea, and shivering (rigors). Severe cases often evolve rapidly, progressing to respiratory distress and requiring intensive care [5]. |
| Middle East Respiratory Syndrome (MERS) | Disease caused by the virus Middle East Respiratory Syndrome Coronavirus (MERS-CoV). Typical symptoms include fever, cough, and shortness of breath. Pneumonia and gastrointestinal symptoms are also commonly reported [14]. |
| Influenza-Like Illness (ILI) | WHO case definitions for ILI have changed many times [15]. As no year of publication will be settled to screen for studies in our review, we will adopt the most reported, which are:<br>• WHO 1999: “a sudden onset of fever, a temperature $>38^{\circ}\text{C}$ and a cough or sore throat in the absence of another diagnosis”.<br>WHO 2018: “an acute respiratory illness with measured temperature of $\geq 38^{\circ}\text{C}$ and cough, with onset within 10 days” [15]. |
| Respiratory Syncytial Virus (RSV)-related ARI | RSV is an RNA virus, classified as a pneumovirus. Is considered the most common cause of lower respiratory tract illness in young infants. The illness commonly manifest are bronchiolitis and pneumonia. These illnesses typically begin with upper respiratory symptoms and fever, then progress over several days to dyspnea, cough, wheezing, and/or crackles on chest auscultation [16]. |
| Parainfluenza Virus type III (PIV3)-related ARI | Most frequently among young infants and immunocompromised children and adults. This type of infection may present pneumonia and/or bronchiolitis clinical syndromes [17]. |
| 2019 GNI per capita between \$4,046 and \$12,535 [18] | Albania; American Samoa; Argentina; Armenia; Azerbaijan; Belarus; Belize; Bosnia and Herzegovina; Botswana; Brazil; Bulgaria; China; Colombia; Costa Rica; Cuba; Dominica; Dominican Republic; Ecuador; Equatorial Guinea; Fiji; Gabon; Georgia; Grenada; Guatemala; Guyana; Indonesia; Iran, Islamic Rep.; Iraq; Jamaica; Jordan; Kazakhstan; Kosovo; Lebanon; Libya; Malaysia; Maldives; Marshall Islands; Mexico; Montenegro; Namibia; North Macedonia; Paraguay; Peru; Russian Federation; Samoa; Serbia; South Africa; St. Lucia; St. Vincent and the Grenadines; Suriname; Thailand; Tonga; Turkey; Turkmenistan; Tuvalu; Venezuela, RB [18]. |

ARI: Acute Respiratory Infections; GNI: Gross National Income; WHO: World Health Organization.

For the economic evidence set, we will evaluate the economic perspective, study scope, and study design of manuscripts for inclusion. From an economic perspective, we will accept individual, provider, societal, or third-party payers. For study scope, reports that estimate costs at institutional, municipal, regional, or national level will be included for analysis [7]. Multi-country studies (international scope) will be included only if they provide an explicit, distinguished, and detailed economic estimate for one of the UMIC investigated (Table 1).

Considering study design, we will include partial economic evaluation, such as COI studies and burden of illness/diseases, database analysis, observational reports (cross-sectional studies, and prospective and retrospective cohort), and economic modelling studies [7]. Due to the diversity of geographical settings that will be investigated, and aiming for a comparative assessment of costs analysis, we will only include studies that present cost components (Table 1) in disjointed analysis. There will be no restrictions on the time horizon for the studies, language, year of publication, or publication status.

We will exclude letters, reviews, commentaries, editorials, case reports, case series, and papers without sufficient information to clearly identify methods, sources, or unit costs. Complete economic evaluations studies, such as cost-effectiveness, cost-utility, cost-benefit, and cost-minimization analysis will also be excluded. Moreover, clinical trials of interventions with economic evidence, and studies that presented proposals for new prevention measures, assuming the cost that could generate, will not be included.

Our exclusion criteria will also be applied to studies that measure and report alone intangible costs, indirect costs, or societal economic perspective (Table 1), due to the difficulties of accurately quantifying certain costs and comparing social and cultural characteristics between different nations. In like manner, we will exclude epidemiological studies using disability-adjusted life years as result [7]. In view of our goal to identify the economic impact of the history of disease, cost analysis that estimate financial burden on government assistance programs during a health crisis, such as COVID-19 pandemic, will also be excluded.

Conference and journals abstract that meet the eligible criteria will be directly included if a complete extraction of data and quality analysis can be performed. In case of insufficient data, we will contact the authors of involved study to solicit relevant information. A period of three months for response will be established. If there is no return or any impossibility within the contact, the study will be included into the review and filed in “Studies Awaiting Classification”. The same label will be applied to non-English language reports from which it is not possible to perform a valid translation and, therefore, the systematic evaluation [19].

For the viral ARI set, we will adopt the definitions presented by the WHO guidelines for infection prevention and control of epidemic- and pandemic-prone ARI in health care [2]. Thus, we will search for diseases that cause acute respiratory tract infection in humans, including pneumonia and/or acute respiratory distress syndrome (e.g. bronchiolitis); cause severe complications in susceptible individuals with apparently normal immune defense systems; are caused by a viral microorganism; and may constitute a public health emergency of international concern. Therefore, the following viral ARI will be included: COVID-19, SARS, Middle East Respiratory Syndrome (MERS), Influenza-Like Illness (ILI), Respiratory Syncytial Virus (RSV)-related ARI, and Parainfluenza Virus type III (PIV3)-related ARI. The definitions adopted to characterize the diseases are presented in Table 1.

Studies conducted on other types of viral ARI than established in our review (Table 1), such as laryngitis, pharyngitis, rhinitis, viral ARI in non-humans etc., will be excluded. Furthermore, economic evaluations that included one of our viral ARI investigated as secondary infection will be excluded if they do not report the costs in a distinguished manner.

Finally, the UMIC set will follow the classification established by the World Bank, according to 2019 Gross National Income (GNI) per capita. Thus, we will include in our review countries that presented a GNI per capita between $ 4,046 and $ 12,535 [18]. Table 1 details the list of the included countries.

### Search methods for identification of studies

Our search for economic shreds of evidence will be performed in five main databases: the Medical Literature Analysis and Retrieval System Online (MEDLINE); Excerpta Medica database (EMBASE); Latin American and Caribbean Health Sciences Literature (LILACS); Cumulative Index of Nursing and Allied Health Literature (CINAHL), and EconLit. Considering the cost analysis topic of investigation, a more broadly specified search will be performed in The Centre for Review and Dissemination (CRD) Library, incorporating the Database of Abstracts of Reviews of Effects (DARE), the National Health Service (NHS) Economic Evaluation (NHS EED), and NHS Health Technology Assessment (NHS HTA) databases [20].

Moreover, we will also search for evidence in the preprint server for Health Sciences, MedRxiv, and the preprint platform Research Square. Finally, hand searches of the bibliographies of included studies and relevant reviews identified during the screening process will be undertaken to identify any additional relevant study for inclusion in our review [10].

The search strategy will be developed with the guidance of an expert research librarian (*MESP*). The sensible structure will include and combine the three main “sets” of interest of this review: economic evidence, viral ARI, and UMIC. For the economic evidence search strategy, we will adapt the search filters provided by Shemilt and collaborators [20] in the Cochrane Handbook, and combine strategies developed by other authors in previously systematic reviews of economic burden [4,11]. For the viral ARI set, the search strategy will be based on controlled vocabulary for each disease. Lastly, UMIC search terms will follow the list of the World Bank [18] and controlled vocabulary related to developing countries [4,11]. No year of publication filter will be applied.

The search strategy that will be applied to MEDLINE (via PubMed) is shown in Table 2. Adaptations of this search strategy will be taken across the different databases. A detailed strategy for each of the databases will be published in the final review.

**Table 2.**
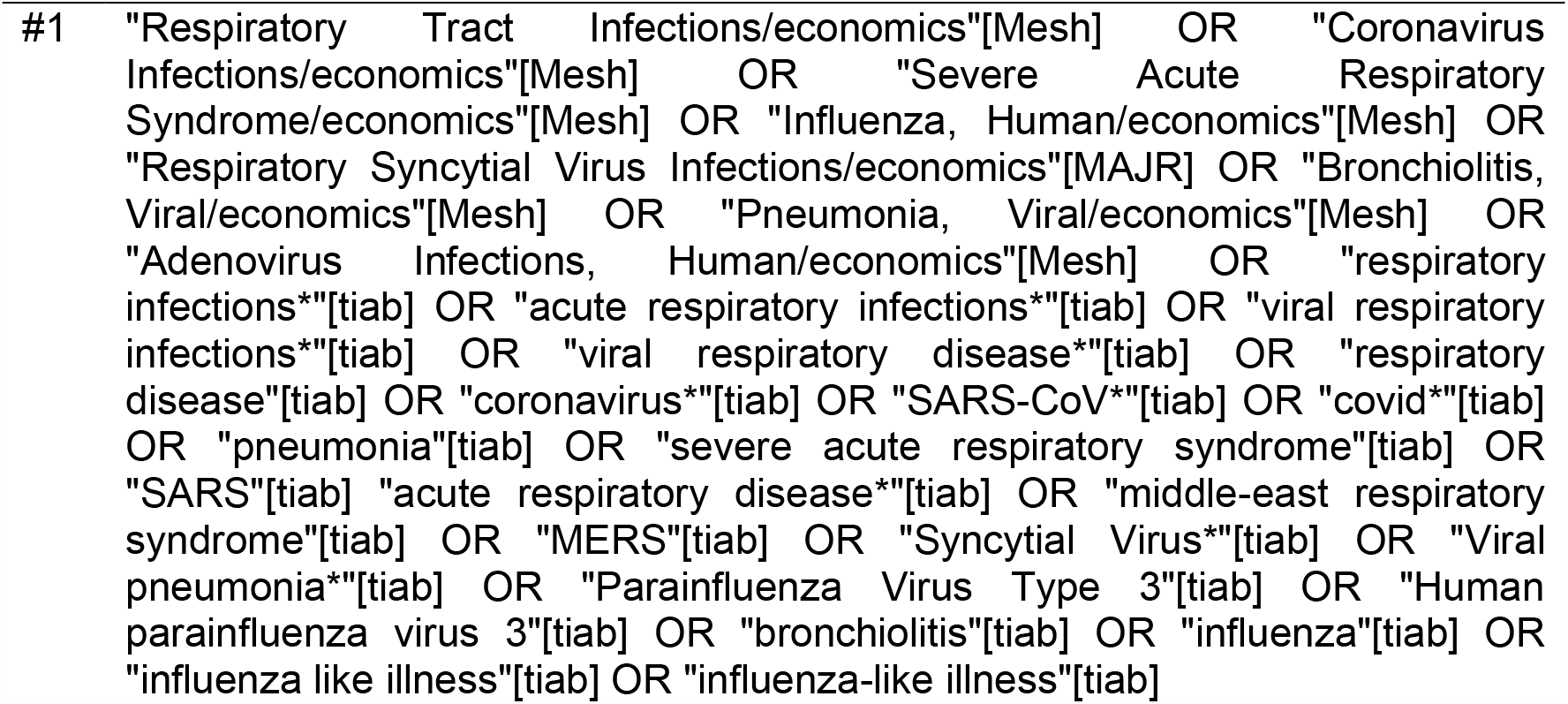
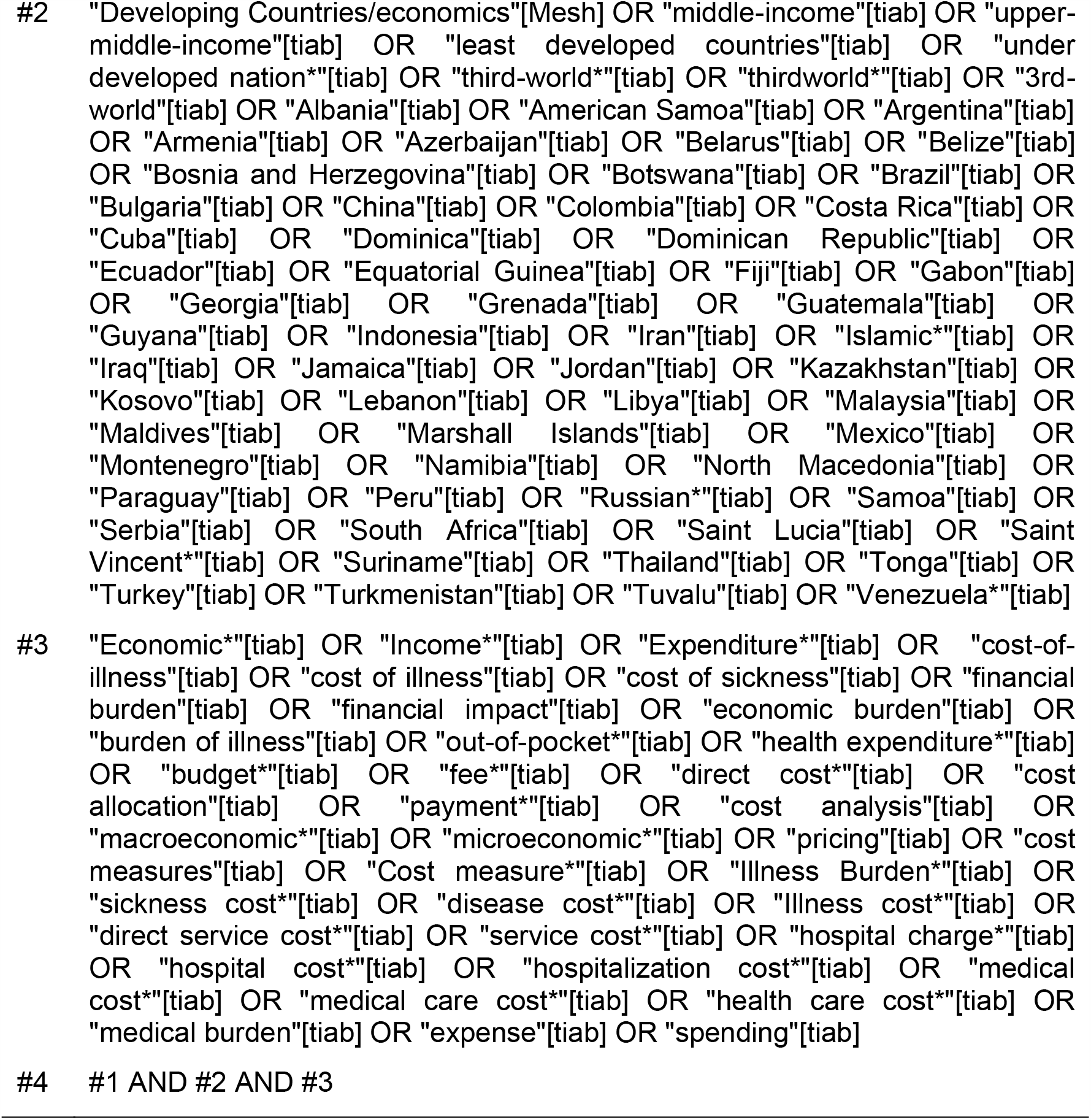
Search strategy for MEDLINE (via PubMed).

### Selection of studies

The reports identified through our search strategy will be uploaded to the web application Rayyan (http://rayyan.qcri.org) [21]. After removing duplicates, two review authors (*CRRF and one of ACPNP, APR, FSAR, GFMT or GSR*) will independently screen titles and abstracts of all recovered studies against pre-established eligibility criteria. The selected studies will then be fully read, by the same review authors, for further scrutiny. We will exclude full-text articles that do not meet the eligibility criteria and then present reasons for exclusions in the final review. At all stages, disagreements between reviewers will be discussed and resolved consensually. If necessary, the opinion of a third reviewer (*MMCL or VFMT*) will be consulted.

If a same study, using the same sample and methods, is found in different published reports (e.g. abstract in congress, study protocol, and full-text article), all of them will be included, but analyzed as a unique study. If different studies in a same population, with the same viral ARI investigated, are found completing overlapping time horizons, the report with the smaller period of analysis will be excluded. If a similar situation occurs without the temporal superposition, both reports will be included and analyzed as different studies.

### Data extraction and management

For data extraction, we will follow a process of two stages. Firstly, two review authors (*APR and one of ACPNP, FSAR, GFMT or GSR*) will extract and compile the data of each included study into a prestructured and piloted data collection form, designed specifically for this synthesis. Then, the completed forms will be reviewed by a third review author (*CRRF*) for accuracy and completeness. Discrepancies will be resolved by discussion and consensus among the authors.

In the event where there is missing data that we deem important, the corresponding author of the study will be contacted. The process will follow as previously presented for conferences and journals abstract inclusion. If the study fulfills all eligibility criteria but we receive no return within the contact, the label “Study Awaiting Classification” will be applied [19].

An initial version of the data collection form will be structured combining general information about the included studies, and elements of the three main “sets” of interest in our review [22]. More specifically, in this version we will extract information on: i) study identification (first author and year); ii) type of viral ARI investigated; iii) geographical setting (UMIC); iv) study design; v) study size (e.g. population-based studies or sampled-based studies); vi) population characteristics (e.g. age and gender); vii) epidemiological approach (prevalent or incident); viii) study scope (data source); ix) economic perspective; x) study duration/time horizon; xi) cost components; xii) total cost; xiii) cost conversion (see “standardization of cost data”); and xiv) conflicts of interest. Table 1 details the elements of each category and presents definitions.

We will pilot this initial version on a randomly selected sample of included studies (maximum five, if available), and then determine the final version. Modifications will be reported in the systematic review publication.

#### Standardization of cost data

To reach a better comparability between the different currencies, all reported costs in the included studies will be converted to Purchasing Power Parity (PPP)-adjusted US$ 2021 (Int$ 2021) [20]. For this, we will use the free online cost converter tool from Campbell and Cochrane Economics Methods Group Evidence for Policy and Practice Information and Coordination Centre (CCEMG-EPPI Centre) (https://eppi.ioe.ac.uk/costconversion/) [23].

When the base year of the currency is not reported or cannot be inferred from the manuscript (e.g. last year of data collection), it will be assumed as reference year the year before the publication of the manuscript. For studies presenting estimates for more than one year, the most recent estimate will be extracted. No adjustment will be made to economic outcomes expressed as ratios or percentages, e.g. percentage of Gross Domestic Product (GDP) [4].

### Quality assessment

After extraction, the quality of included studies will be appraised independently by two review authors (*CRRF and MMCL*). Discrepancies will be solved by consensus and, if necessary, a third review author will be consulted (*ANA or VFMT*).

As previously reported, to the best of our knowledge, there is currently no formal guideline or consensus on recommended tools or checklists to assess the quality of studies that report the economic burden of diseases [4,7,9]. In our review, we will follow the checklist used by Oliveira, Itria, and Lima [9], that was based on the protocol proposed by the British Medical Journal (BMJ) [24], and on the Consolidated Health Economic Evaluation Reporting Standards (CHEERS), version 2013, developed by the International Society for Pharmacology and Outcomes Research (ISPOR) [25].

The checklist is structured in three blocks of questions, totalizing 29 items that aim to characterize a health cost analysis. The first block contains 11 items referred to the study design. The second block analyzes the data collection process of each study in 9 items. Lastly, the third block performs the analysis and interpretation of the results in 9 items. Each item is rated in three grades: “yes”, “no”, and “not applicable”. At the end of the classification, a relative value of each of these grades is settled for each study. The goal is to check the percentages of “yes” for each question [9].

### Summary measures and synthesis of results

Due to the expected heterogeneity of the investigated topic in terms of design, study object, setting, methodological perspective and other characteristics, a quantitative synthesis of the burden estimates is unlikely to be appropriate. Thus, the results of included studies will be interpreted qualitatively in a narrative synthesis.

The main aspect of aggregation will be the type of viral ARI: COVID-19, SARS, MERS, ILI, RSV-related ARI, or PIV3-related ARI. We will then focus on the overall economic burden of the diseases on health systems and population; total direct cost; the contribution of different cost components to the economic burden (e.g. pharmacological therapy, hospitalization); comparative assessments of costs analysis across geographical location and time horizon; and current research gaps. Moreover, we intend to identify, when presented, prevalence and incidence rates of each disease. We will follow the PRISMA statement and checklist to undertake and report the findings of our systematic review [13].

## Discussion and study limitations

There are some goals that we believe that our systematic review may achieve. Firstly, we will present the economic burden viral ARI to health systems, patients, and even society. If possible, we would like to report how these cost impacts on GDP of countries, aiming to present data to support the development of programs to respond to viral ARI.

When comparing and identifying major costs drives related to a viral ARI in different countries with sociodemographic similarities, we expect to point out perspectives and actions that may or may not have worked successfully. The same expectation is applicable to analyze different time horizons in the same country. These lessons may inform decision-makers and researchers to support formulation and refinement of policy and research objectives in viral ARI.

We anticipate some limitations in our study related to bias in viral ARI reporting and estimating burden of the diseases. As COVID-19 is an emerging public health concern, with many topics still under discovery, we may encounter a lack of studies or incomplete analysis with dubious results. We intend to overcome these limitations with a sensible analysis of the quality and relevance of included studies, and a further update of this systematic review two years after the first publication.

Finally, we intend to share our findings in conferences, peer-review journals, and social media platforms.

### Support for the review

This study has the support of the Coordenaçã o de Aperfeiçoamento de Pessoal de Ní vel Superior – Brasil (CAPES).

## Data Availability

All data referred to in the manuscript are available.

